# A Wearable In-pad Diagnostic for the Detection of Disease Biomarkers in Menstruation Blood

**DOI:** 10.1101/2024.03.22.24304704

**Authors:** Lucas Dosnon, Thomas Rduch, Charlotte Meyer, Inge K. Herrmann

## Abstract

The pain-free regular monitoring of blood-based biomarkers is a highly appealing yet difficult-to-realize approach for the early detection of pathological changes, including cancers, infections, or metabolic diseases, such as diabetes. While a major focus of the research community lies on the investigation of pain-free blood sampling and devices for venous blood analysis, menstruation blood remains a largely ignored sampling source. Growing evidence shows excellent correlation between biomarker levels in menstruation blood and venous blood for an entire clinical panel of analytes. Here, we introduce a wearable, microfluidic diagnostic platform integrated into standard hygiene pads for the electronic-free naked eye-readable direct detection of disease biomarkers in menstruation blood (MenstruAI). We demonstrate semi-quantitative biomarker detection from menstruation using infection and inflammation biomarker C-reactive protein (CRP), gynecological cancer biomarkers (CEA and CA-125), and endometriosis biomarker CA-125 as representative examples of relevant proteinaceous biomarkers. The color-changes induced by the presence of these biomarkers can be read-out by the naked eye as well as by a machine-learning algorithm implemented into a smartphone-app, enabling semi-quantitative analysis. The presented MenstruAI platform has the potential to revolutionize women’s health by providing a non-invasive, affordable, and accessible approach to health monitoring, potentially democratizing healthcare by making health services more available and equitable.

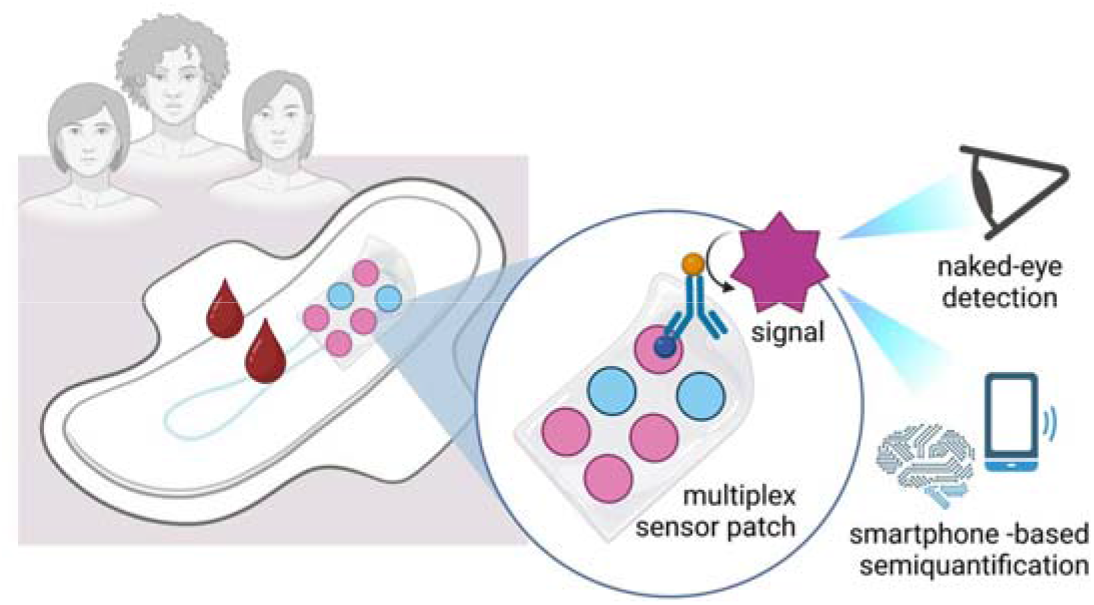

## INTRODUCTION

In modern medicine, blood tests and blood panels are indispensable, providing valuable insights into various physiological aspects to make informed decisions about patient care. The levels of molecular constituents in blood are directly associated with the physiological state of the body.^1–4^ Traditionally, blood testing has been confined to laboratory environments, necessitated by the requirement for sample pre-processing, intricate analytical methods, and specialized instrumentation.^5^ However, the diagnostic significance of blood renders it a prime candidate for point-of-care (POC) diagnostic applications.^6^ While lateral flow assay (LFA)-based analyses of saliva or urine are well-established, the analysis of whole blood samples, which contain many more biomarkers, typically requires invasive blood collection as well as intricate sample preprocessing.^7^ Capillary blood samples obtained through finger pricks are most frequently used.^8^ Nonetheless, blood collection is invasive, leading to poor compliance and reluctant adoption by both patients and, even more so, seemingly healthy individuals. Additionally, using blood in point-of-care devices presents increased barriers to reproducibility and ease of use due to coagulation that hinders its usage, or simply the interference from the red color that may alter a colorimetric readout on a paper-based sensor.^9^ As a result, most of the research conducted on blood-based point-of-care devices involves the use of processed blood. This includes blood containing anticoagulants, blood that has been diluted beforehand with a running buffer, or blood that has been separated from certain components, such as red blood cells, to use only the remaining plasma.^10–13^ These intricate processing steps pose a significant barrier to regular, low-cost health monitoring based on blood biomarkers. While a major focus of the research community lies in investigating pain-free blood sampling and devices for venous blood analysis, menstrual blood appears to be a largely ignored sampling source. Even though every month, 1.8 billion people menstruate,^14^ menstruation blood is largely under-represented in both basic and translation research.^15^ Interestingly, menstrual blood has shown a considerable correlation with systemic blood in terms of protein content through molecular proteomic analysis. Additionally, it contains 385 unique proteins that open the door to further analysis and understanding of women’s health.^16,17^ As a result, menstruation blood is a promising alternative for the noninvasive collection of blood for diagnosis and health monitoring for the menstruating population. Such an approach may be especially promising and economically viable for resource-constrained settings where patients have no access to regular check-ups, as well as diseases with a relatively low prevalence in the population, where preventative testing at the doctor’s office is not economically feasible.^18^ In addition to providing a non-invasive means of accessing blood, menstrual blood is characterized by lower concentrations of coagulation factors, along with reduced levels of hemoglobin and hematocrit.^19^ These properties render menstrual blood particularly well-suited for point-of-care testing, circumventing the need for the preprocessing steps often necessary in the analysis of venous blood. Recent studies have explored the utilization of menstrual blood for health monitoring purposes, focusing on aspects such as volume control of blood loss^20^ and pH levels.^21^ However, these investigations have not leveraged menstrual blood for gaining valuable insights into the physiological state of an individual.

In this work, we report the design and development of a non-invasive low-cost paper based diagnostic platform for the effortless direct on-pad detection of disease biomarkers in menstruation blood by naked-eye detection or smartphone-app (termed MenstruAI), not requiring pre- or post-processing. We demonstrate direct semi-quantitative biomarker detection from menstruation blood using infection and inflammation biomarker C-reactive protein (CRP), gynecological cancer biomarkers (CEA and CA-125), and endometriosis biomarker CA-125 as representative examples of women’s health-relevant proteinaceous biomarkers. The versatile, multiplex detection platform is integrated into a standard hygiene pad. This first-of-its-kind point-of-care device relies on a design that allows both fluidic manipulation and multiplexing for an electronic-free readout, enabling completely passive (not involving human intervention other than deciding to use MenstruAI pads) and reproducible biomarker measurements. The MenstruAI platform directly promotes the use of a sample often seen as waste and increases the accessibility of women to cutting-edge technologies centered on women’s health. It may signal to the user that additional checks at a doctor’s office may be advisable, and with it have transformative impact on women’s health by offering a non-invasive, cost-effective, and barrier-free route to close-meshed health monitoring.

**Figure 1:**
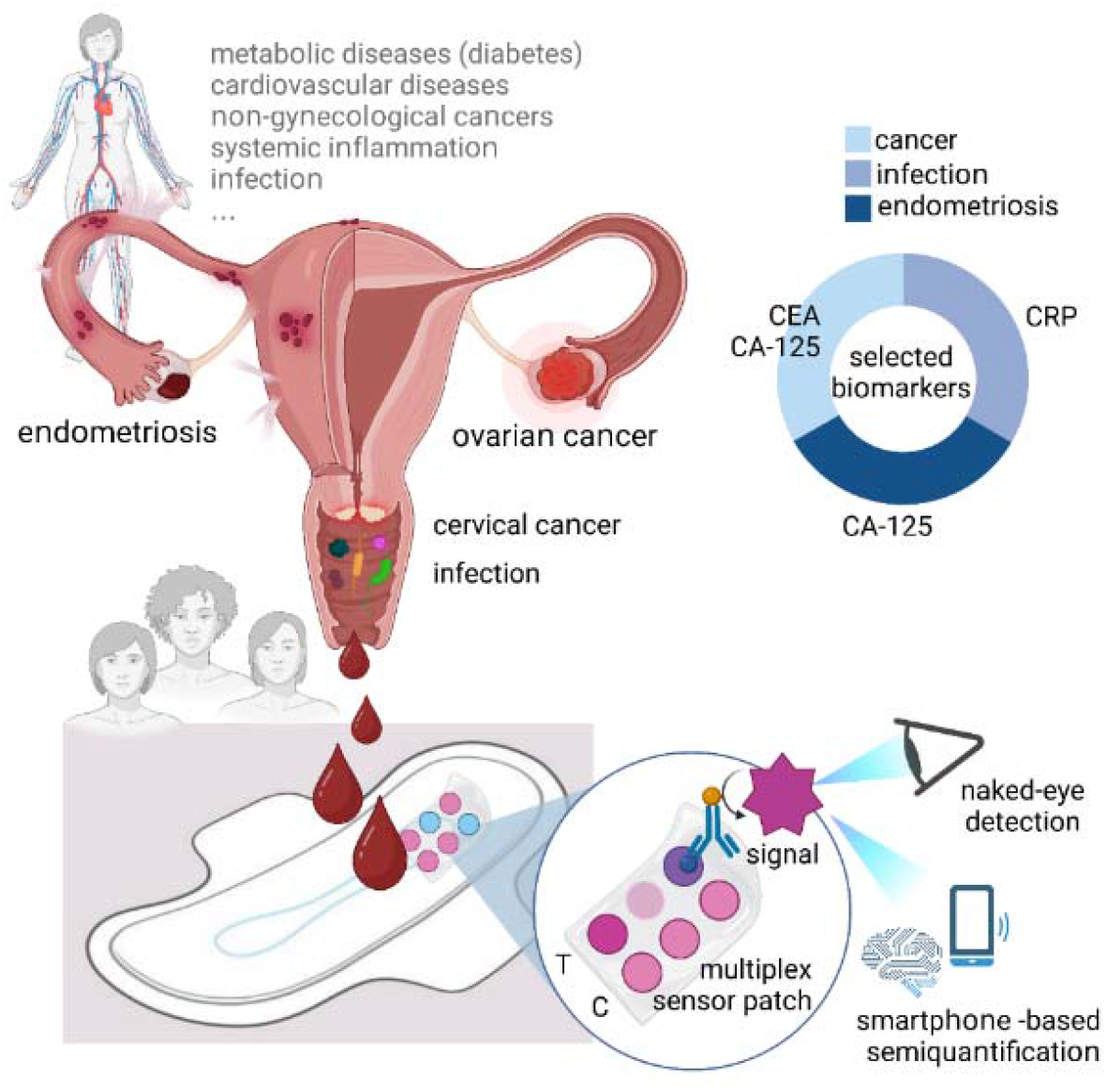
Concept of menstruation blood biosensor integrated into hygiene pads enabling direct naked-eye readable semi-quantitative analysis of biomarkers in unprocessed menstruation blood. The platform termed MenstruAI is designed to include infection/inflammation, cancer and endometriosis biomarkers based on an in-pad integrated lateral flow assay (LFA) platform with integrated volume control.

## RESULTS & DISCUSSION

### Establishment of LFA for CEA, CA-125 and CRP in serum and whole blood

To address the unmet need for affordable and accessible women’s health diagnostics, we sought to design a wearable for the reliable, multiplex detection and semi-quantification of health-relevant analytes in menstrual blood. The solution is designed to provide an accessible, cost-effective, reproducible, non-invasive method for detecting important health biomarkers through menstrual blood, offering a convenient, comfortable, and accessible solution for personal health monitoring to the menstruating population.

First, lateral flow assay (LFA) sensors were designed for the detection and semi-quantification of individual biomarkers in relevant biofluids (including whole blood) and clinically relevant detection windows. CEA, CA-125 and CRP were selected as representative biomarkers, covering diagnostically interesting areas of cancer, endometriosis, and infection, making them particularly relevant for women’s health. Additionally, the presence of these biomarkers is indicative of a pathology since baseline levels in healthy individuals are typically low and in the region of the analytical detection limits of a typical LFA. Careful optimization of the LFA sensors was performed to achieve detection and semi-quantification in i) human serum, and ii) unprocessed human whole blood for practicality, ease of use and to eliminate the need for sample transformation or addition of a running buffer. The LFAs were engineered to incorporate both a test line, a control line and if needed, an antigen line to circumvent to so-called hook effect.^22^ Antibody-functionalized gold nanoparticles were used as recognition element for the different analytes. For assay initiation, a defined volume of the fluid sample (i.e. serum or whole blood) was applied to the sample pad. This sample subsequently migrated towards the conjugate pad, where the target proteins within the sample interacted with functionalized gold nanoparticles pre-deposited on the conjugate pad via antigen-antibody reaction. The test line on the nitrocellulose (NC) membranes consisted of antibodies designed to capture these protein-nanoparticle complexes, thus facilitating a sandwich reaction that resulted in the visualization of the test line. After demonstrating feasibility in serum, the assay was adapted for use with unprocessed human whole blood. A Fusion 5 blood separation membrane was introduced to effectively filter out red blood cells, successfully retaining them within the glass fibers of the sample separation pad. This selective filtration ensured that only plasma progressed through the assay. Consequently, the red hue typically associated with blood was removed, allowing for a clear and unaltered measurement of color intensity at the test and control lines.

### Direct measurement of CEA level in serum and whole blood

CEA was selected as first candidate biomarker relevant in many malignant diseases. Often naturally present at concentration below 5.0 ng/mL in healthy individuals, it can be used to indicate the presence of tumors or spreading cancer when reaching values above 20 ng/mL.^23,24^ Thus, a gold-based LFA for CEA in the clinically relevant detection window (0-500 ng/mL) was established using human-CEA antibodies immobilized on 40 nm spherical gold nanoparticles. In human serum, a logarithmic response curve was observed with increasing concentration of CEA between 0 and 500 ng/mL (Fig S1). A linear response was obtained between 0 and 50 ng/mL with a linear correlation coefficient R^2^ of 0.94 (Fig 2a). For human whole blood, a linear response was obtained on the entire range of concentration, between 0 and 500 ng/mL with a coefficient R^2^ of 0.94 (Fig 2b). Taken together, this assay configuration demonstrated a linear response in the diagnostically important concentration range and sufficient robustness for reliable detection of CEA in a diversity of samples, including serum, plasma, and unprocessed human whole blood.

**Figure 2:**
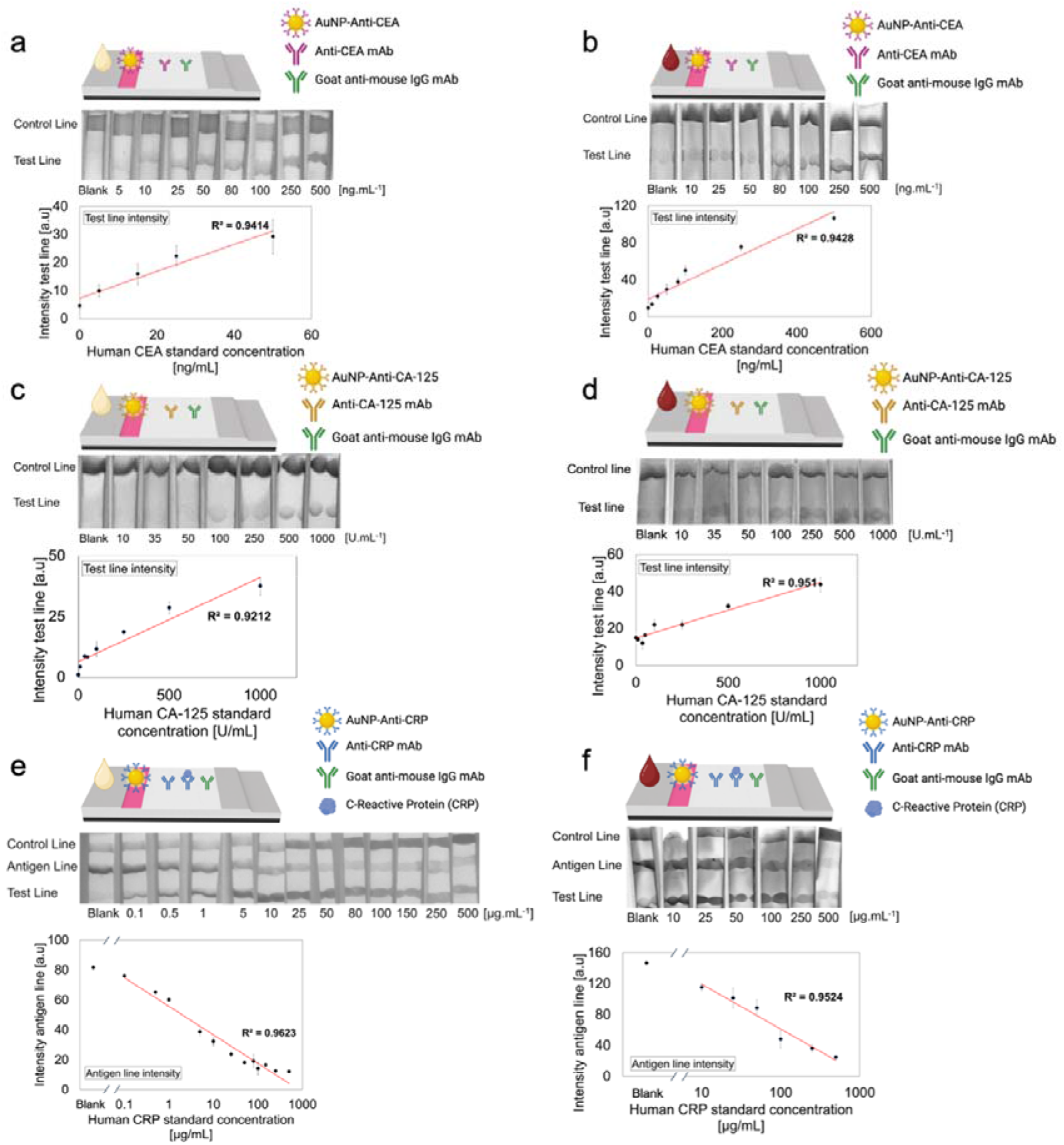
Design of LFAs suitable for biomarker detection in human serum and whole blood in clinically relevant detection windows. a) Design of CEA LFA assay using human CEA-antibody functionalized 40 nm gold nanoparticles in human serum and b) in fresh, unprocessed human whole blood. c) Design of CA-125 LFA assay using human CA-125-antibody functionalized 40 nm gold nanoparticles in human serum and d) in fresh, unprocessed human whole blood. e) Design of CRP LFA assay using human CRP-antibody functionalized 20 nm gold nanoparticles in human serum and f) in fresh, unprocessed human whole blood. Assays were performed in triplicate using independent blood donors (N=3).

### Direct measurement of CA-125 level in serum and whole blood

In addition to CEA, CA-125 was targeted due to its importance in gynecological cancers and endometriosis diagnostics. The conventional upper limit of normal CA-125 in serum is 35 U/mL.^25,26^ Studies^27^ showed that 1% of healthy women and 82% of patients with ovarian cancer had CA-125 levels above this limit. Additionally, women suffering from endometriosis have been shown to have an elevated level of CA-125, reaching values >100 U/mL.^28^ The clinical range of interest of CA-125 is relatively large, with values reaching 1000 U/mL and higher. Therefore, a gold based LFA in the clinically relevant detection window (0-1000 U/mL) was established. Human CA-125 antibodies were immobilized on 40 nm spherical gold nanoparticles. In human serum, a linear response was obtained over the range between 0 and 1000 U/mL with a coefficient R^2^ of 0.92 (Fig 2c). When using human whole blood, the linear response was even more satisfying reaching a coefficient R^2^ of 0.95 (Fig 2d). This demonstrates the capacity of the LFA to perform CA-125 detection over the chosen diagnostically relevant range in a both serum and whole blood.

### Direct measurement of CRP level in serum and whole blood

Finally, CRP was selected as an inflammation biomarker widely used in clinical settings to measure acute inflammation,^29^ but also more recently to assess the risk of developing cardiovascular disease.^30^ CRP is very often used as a model analyte for LFA. However, it is rarely measurable over the full CRP concentration range (0 to 500 µg/mL) due to the so-called “hook effect”.^22^ The hook effect is a known effect in immunoassays that occurs when high concentrations of analytes lead to decreased signal intensity due to the saturation of binding sites on the test line. The formation of the antigen-antibody complex is prevented, resulting in a false-negatives. We designed a three lines LFA geometry, incorporating a sandwich assay-based line that allows to perform the detection of CRP between 0 and 10 µg/mL (Fig S2a, b, c). Additionally, a competitive assay-based line with the pre-immobilized antigen was added, to counter the hook effect that takes place for the detection of CRP level at concentrations above 10 µg/mL (Fig. S2d). The competitive assay-based line comprises of the antigen already bound to pre-immobilized CRP antibody. Thus, in the presence of low concentrations of CRP in the sample, this line has the highest signal intensity, as most AuNP conjugates will get immobilized on it. At high concentrations of CRP, most of the proteins will bind to the AuNP conjugates, which will therefore not bind to the antigen deposited on the line leading to a gradual decrease of the signal intensity (Fig 2e, f). In both human serum and human whole blood, introduction of an antigen line led to a very satisfying linear detection range of CRP throughout a clinically relevant window with R^2^ coefficients of 0.96 (0.1-500 µg/mL) and 0.95 (10-500 µg/mL), respectively (Fig 2e, f). This three lines LFA geometry therefore enabled the detection of CRP throughout the full concentration range (0 to 500 µg/mL).

### Design of soft-silicon embedded paper-based wearable sensors with integrated volume control

Leveraging the developed LFA sensing technology, we investigated design strategies for its seamless integration into sanitary products. This integration relies on embedding LFA biosensors into hygiene pads, enabling effortless direct, on-pad analysis of menstrual blood. This approach is guided by several critical design principles, including the simplification of processing steps prior to readout, ensuring minimal procedural complexity. The device’s hallmark is its ability to operate autonomously, necessitating no more than a simple selection of the appropriate pad by the user, thanks to the fully integrated LFA technology.

In line with this principle, our wearable prototypes were specifically engineered to analyze unprocessed human menstrual blood for user-friendly, efficient health monitoring. However, this ambition comes with challenges, notably in ensuring consistent, reproducible sampling amidst the dynamic variability of menstrual blood loss, influenced by individual and physiological factors. Excessive blood contact with the test platform could alter or invalidate results and potentially lead to overflow, affecting the test’s usability. In addition to these technical considerations, the integration must meet critical requirements of biocompatibility, comfort, and user-friendliness. To meet the aforementioned design specifications, we designed two different prototypes, relying on distinctly different volume control approaches.

Prototype 1 was designed with a soft-silicon casing that encases an LFA, featuring an inlet reservoir with a time-dissolvable PVP membrane at its bottom, capable of holding 150 µL of blood for a set duration (Fig 3a). While the polyvinylpyrrolidone (PVP) membrane is intact, the blood stays in the reservoir and the LFA test can run to completion. As blood fills the reservoir, it interacts with the PVP membrane, causing it to degrade and eventually fail, a process adjustable through the membrane’s PVP concentration (Fig 3e). When the membrane fails, any additional blood simply passes through the now open reservoir and is absorbed by a layer beneath the casing. This device was integrated into a hygiene pad, positioned under a polypropylene (PP) transfer layer that directed blood into the inlet reservoir. After the test, the PP layer was removed to reveal the LFA’s readout zone through the transparent silicon casing. This design enables reliable volume control and straightforward readout of the integrated LFA test. The inlet reservoir’s dimensions are adaptable, aligning with the needs of the LFA and ensuring the correct blood volume is used for the test. The modular design effectively manages sample volume, preventing overflow and protecting the test within a clear casing.

**Figure 3:**
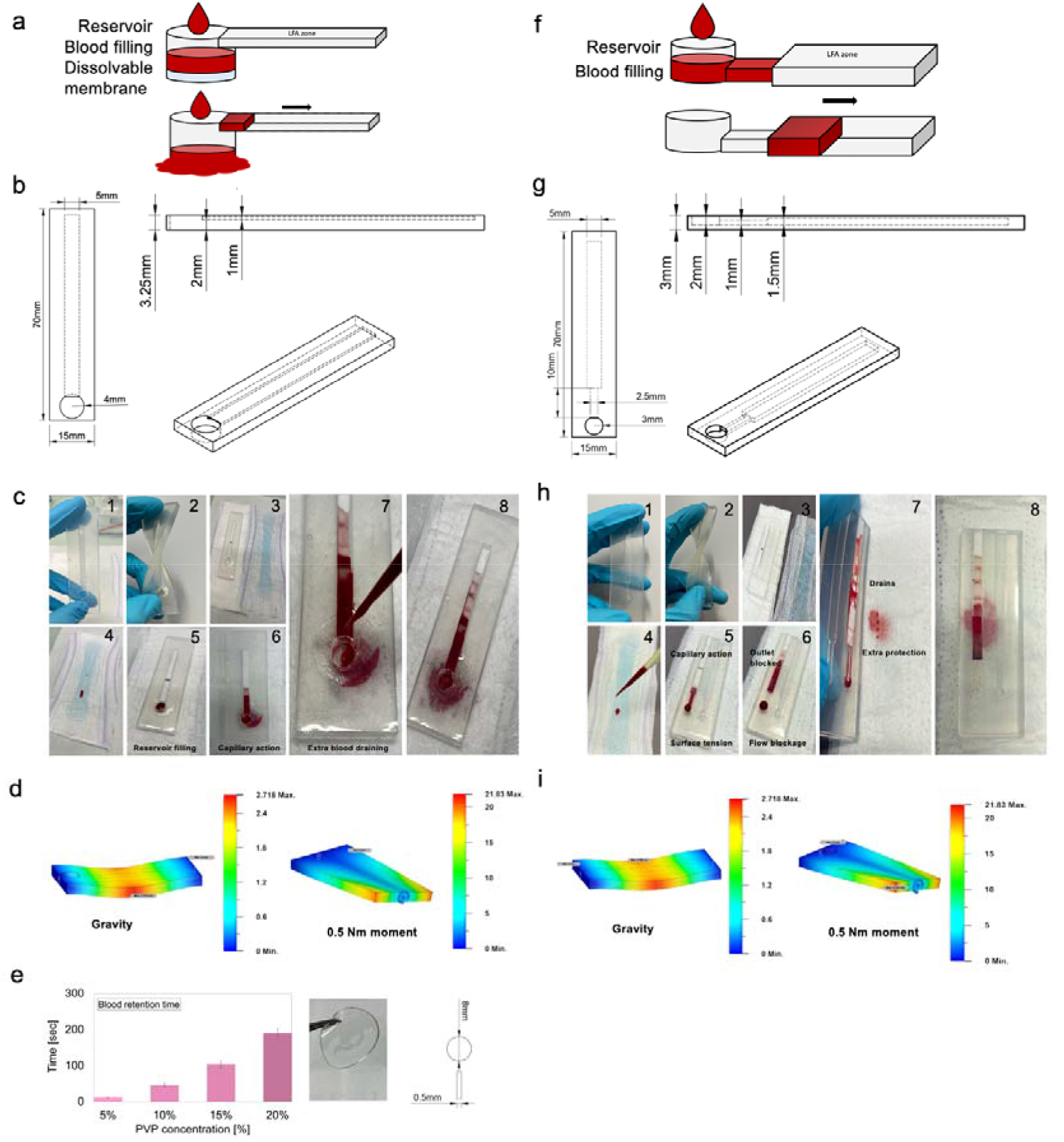
a) Concept and b) technical characteristics of prototype design 1 of soft-silicon embedded wearable paper-based sensor. The design includes a reservoir of known volume with a dissolvable membrane acting as a drain after test completion. c) Integration of prototype 1 embedded into a sanitary pad and demonstration of functional volume control (preventing test overflow) (1,2: soft-silicon casing for paper-based sensor embedding. 3: device placement into sanitary pad. 4: blood collection on first fluid transfer layer. 5,6: capillary-based intake of controlled volume and LFA test completion. 7: draining of additional blood. 8: completed test.) d) Wearability assessed based on mechanical stress response simulations of prototype 1 using gravity and a 0.5 nM moment. e) Dissolvable membrane concept and characteristics showing a controllable dissolving time based on PVP concentration. f) Concept and g) technical characteristics of prototype design 2 of soft-silicon embedded paper-based sensor. The design incorporates an inlet reservoir connected to a capillary channel with predefined dimensions. It utilizes a pressure gradient between the inlet and outlet to precisely control the volume and flow of blood sampled until the correct amount reaches the embedded Lateral Flow Assay (LFA) sensor. h) Integration of prototype 2 embedded into a sanitary pad and demonstration of functional volume control (preventing test overflow) (1,2: soft-silicon casing for paper-based sensor embedding. 3: device placement into sanitary pad. 4: blood collection on first fluid transfer layer. 5,6: capillary-based intake of controlled volume, pressure gradient cancelation and LFA test completion. 7: draining of additional blood. 8: completed test.) i) Wearability assessed based on mechanical stress response simulations of prototype 2 using gravity and a 0.5 nM moment.

Prototype 2 consists of a similar soft silicon-based casing containing the LFA strip. The design of the device is different from prototype 1 as it relies on principles of capillary pressure driven flow microfluidics that allows passive movements of fluids in microchannels (Fig 3f). When a drop of blood is applied on the first PP transfer layer of the hygiene pad, it automatically gets directed above the inlet reservoir that holds a volume of approximately 60 µL. Given the micrometric dimension of the device surface tension forces dominate against gravity. This causes the liquid sample to enter the capillary directing the sample towards the LFA. Inside the microfluidic channel of approximately 25 µL volume, the Laplace pressure at the liquid/air meniscus interface is the driving force responsible for the movement of the fluid:^31^

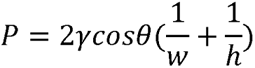

with *γ* denoting the surface tension, *θ* the contact angle, *w* the width and *h* the height of the rectangular channel. With the atmospheric *P*_0_ at the outlet, this generates the right pressure gradient to drive the fluid to the LFA. Additionally, considering an incompressible and non-Newtonian fluid in a laminar flow within a cylindrical capillary, it is possible to use Hagen-Poiseuille equation:^32^

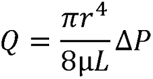

with *Q* the volumetric flow rate, *r,* and *L* the radius and length of the capillary, *µ* the dynamic viscosity of the liquid and Δ*P* the pressure difference between inlet and outlet.

The Washburn equation^33^ then gives the filling rate of a channel with constant capillary pressure:

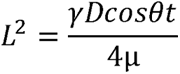

with *L* the length of channel that is filled at time *t, γ* the surface tension, *θ* the contact angle, *D* the diameter of the channel, *µ* the viscosity. This design ensures consistent capillary filling under fixed channel parameters, finely tuned for controlled sample flow to the LFA. Upon contact, the blood is absorbed by the paper-based sensor through capillary action, completing the test. The wetting of the paper causes the LFA’s sample pad within the silicon device to clog, interrupting the air flow between inlet and outlet, and nullifying the pressure gradient, thereby preventing further liquid entry. This mechanism ensures reproducibility and prevents overflow. In addition, we included draining holes beneath the LFA, channeling excess blood into the absorbent layers of the hygiene pad (Fig 3h). For read-out, the top PP layer is easily removable, revealing the LFA’s readout zone through the clear casing. Like Prototype 1, the device’s dimensions and consequently the blood volume are adjustable, offering a flexible solution tailored to the specific design requirements. Finally, the soft silicon utilized in both prototypes demonstrates high flexibility, as validated through comprehensive mechanical stress simulations (Fig3 d, i). Even minor stressors, such as gravity or a mere 0.5 nM moment, induce noticeable deformations. This high malleability ensures that the prototypes are highly responsive and compliant with users’ dynamic movements, making them an ideal choice for wearables designed to accommodate a range of motion.

### Smartphone-assisted semi-quantitative analysis of biomarkers in menstruation blood

With the volume-controlled in-pad LFA platform designed, the final crucial step for an accessible health monitoring platform lies in the reliable readout of the test results. LFA results are often subject to subjective interpretations, leading to frequent false positives or negatives^34,35^ especially when using unprocessed whole blood, which can cause interference such as blood leakage, immobilized red blood cells forming misleading lines, or lysed cells creating a red background. The semi-quantitative output from multiplexed detection of biomarkers like CEA, CA-125, and CRP, including an antigen line in the case of CRP, poses additional challenges for visual evaluation by untrained users, underscoring the need for standardized readout methods.

**Figure 4:**
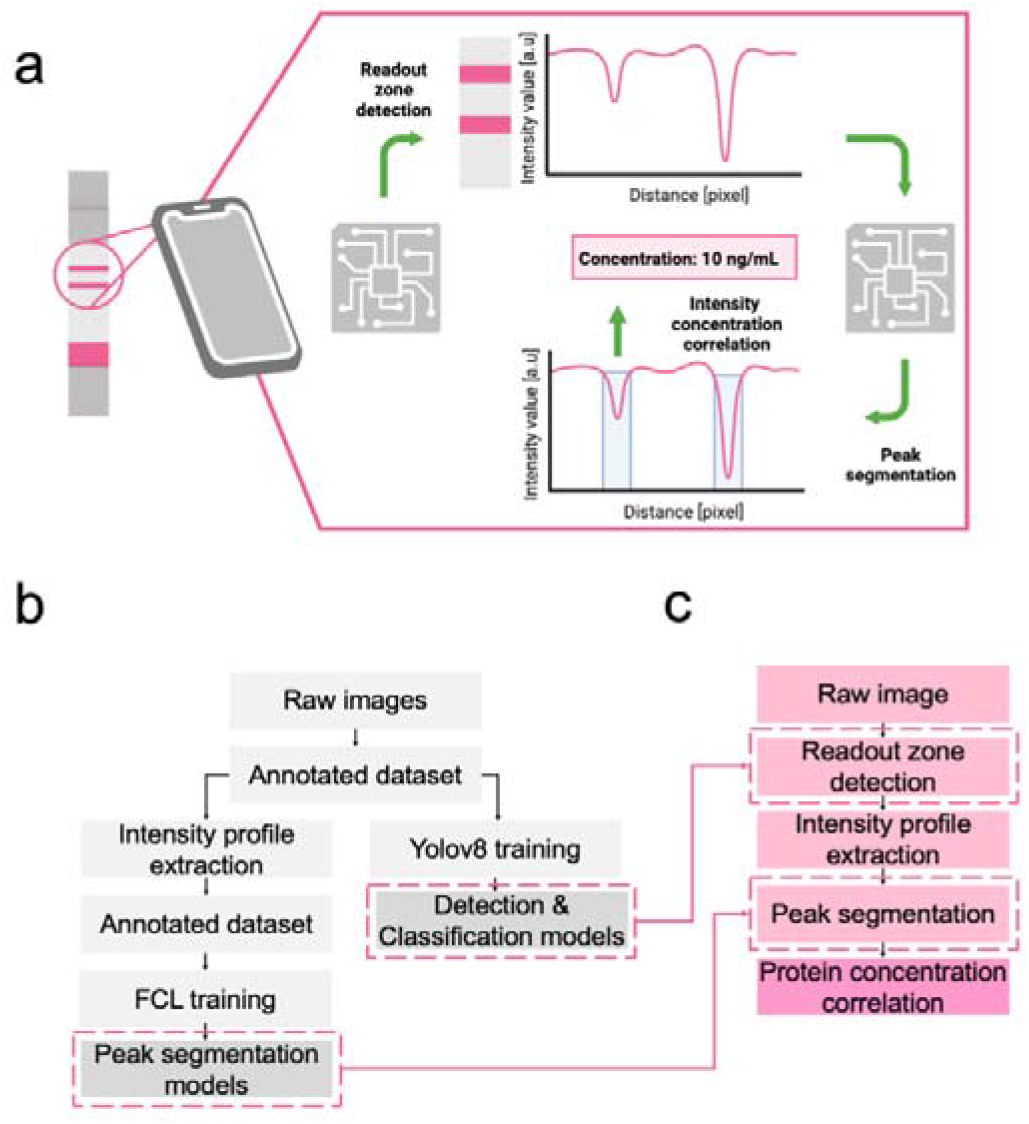
a) Concept of the smartphone-based machine learning (ML) for LFA test analysis. A test image was captured with a smartphone. An ML model was employed to identify the readout zone and classify the result, analyzing the pixel intensity profile to locate peaks, corresponding to control and test line intensities. These intensities can be directly linked to biomarker concentration, based on pre-established integrated calibration. b) Workflow of machine learning model dataset preparation and training. For the training process, a dataset was first generated using a set of raw images of LFA strips, manually labeled. This dataset was then used for the training of two ML models for readout zone detection and classification. Additionally, intensity profiles were extracted from the same images and manually annotated for peak and background regions. This dataset was then used for the training of the fully connected layer (FCL) model to perform automatic peak segmentation on new (unseen) data. c) Illustration of the workflow of the machine learning application. Once the three models were trained, they were employed to detect and classify the readout zone on new LFA strip images. The intensity profiles were then extracted for peak segmentation and biomarker concentration determination.

To address these issues, we developed a fully automatic analysis application using machine learning, deployed on smartphones for standardized assay readouts after pad use. The approach involved using a smartphone-captured image of the LFA strip, and subsequent analysis with segmentation and classification models. The application first automatically detected the readout zone, where colorful lines (control and test lines for CEA and CA-125, and control, test, and antigen lines for CRP LFA) were visible. For this purpose, a pre-trained yolov8 convolutional neural network (CNN) was employed, achieving a box loss of 0.73 and an mAP50 of 0.995 (Fig5 a).

With a confidence threshold of 0.8, it accurately detected the readout zone across the entire test dataset. After segmenting the readout zone, it was processed by a classification CNN and a fully connected layer (FCL) model, enabling peak segmentation on the intensity profile, which displayed 2 or 3 peaks depending on the output type. Without discriminating the output type, the neural network achieved an accuracy, precision, and recall of 0.83, 0.90, and 0.86, respectively. However, when data was classified and separated based on output type, the network’s performance increased, with both models’ accuracies reaching 0.90 and precision over 0.93 (Fig 5b). The application then provides the user with the concentration value of the biomarker measured by the LFA, offering a reliable and immediate interpretation of results (Fig 5d).

**Figure 5:**
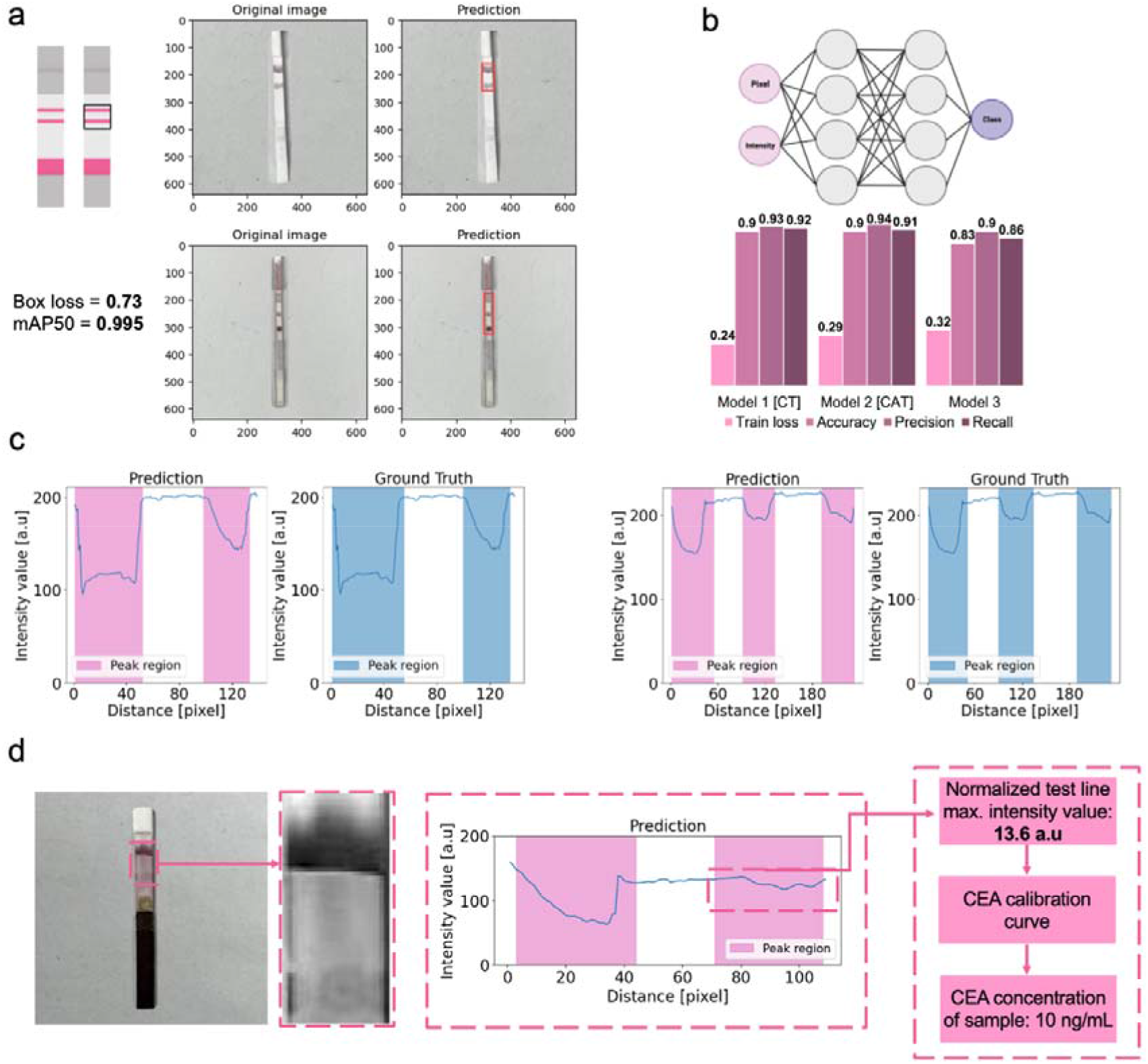
a) Readout zone detection using CNN. The CNN model automatically detected the region of interest where the readout of the test was located. The readout could then be classified. The model reached a box loss of 0.73 and mAP50 of 0.995 after training. b) Concept and performances of FCL models used for peak segmentation. Three models were built similarly with an input layer, hidden layers, and an output layer. Discriminating the output type directly influences positively the performances of the trained models (model 1 and 2) compared to a model trained on all the possible output types (model 3). c) Examples of peak segmentation on intensity profiles displaying 2 and 3 peaks (corresponding to control, test and possibly antigen lines). The FCL model accurately detected the regions corresponding to a peak of intensity on the LFA strips. Comparison between prediction and ground truth demonstrates very satisfactory performances. d) Example use case of automatic CEA quantification from a smartphone-captured image. The readout zone was accurately detected and classified. Peak segmentation was performed by the FCL model, and the value of CEA was obtained after extraction of the test line intensity, which is linked to the biomarker concentration based on pre-established calibration.

Finally, we demonstrate the robust and reliable biomarker detection and semi-quantification directly in unprocessed whole menstruation blood using the developed platform. The obtained menstruation blood was used as a matrix for CRP detection using the designed CRP LFA presented above (Fig 6a-c). In menstruation blood, biomarker quantification results comparable to the detection in human venous whole blood have been obtained, which demonstrate the feasibility of biomarker detection using the envisioned LFA platform directly in unprocessed menstruation blood.

**Figure 6:**
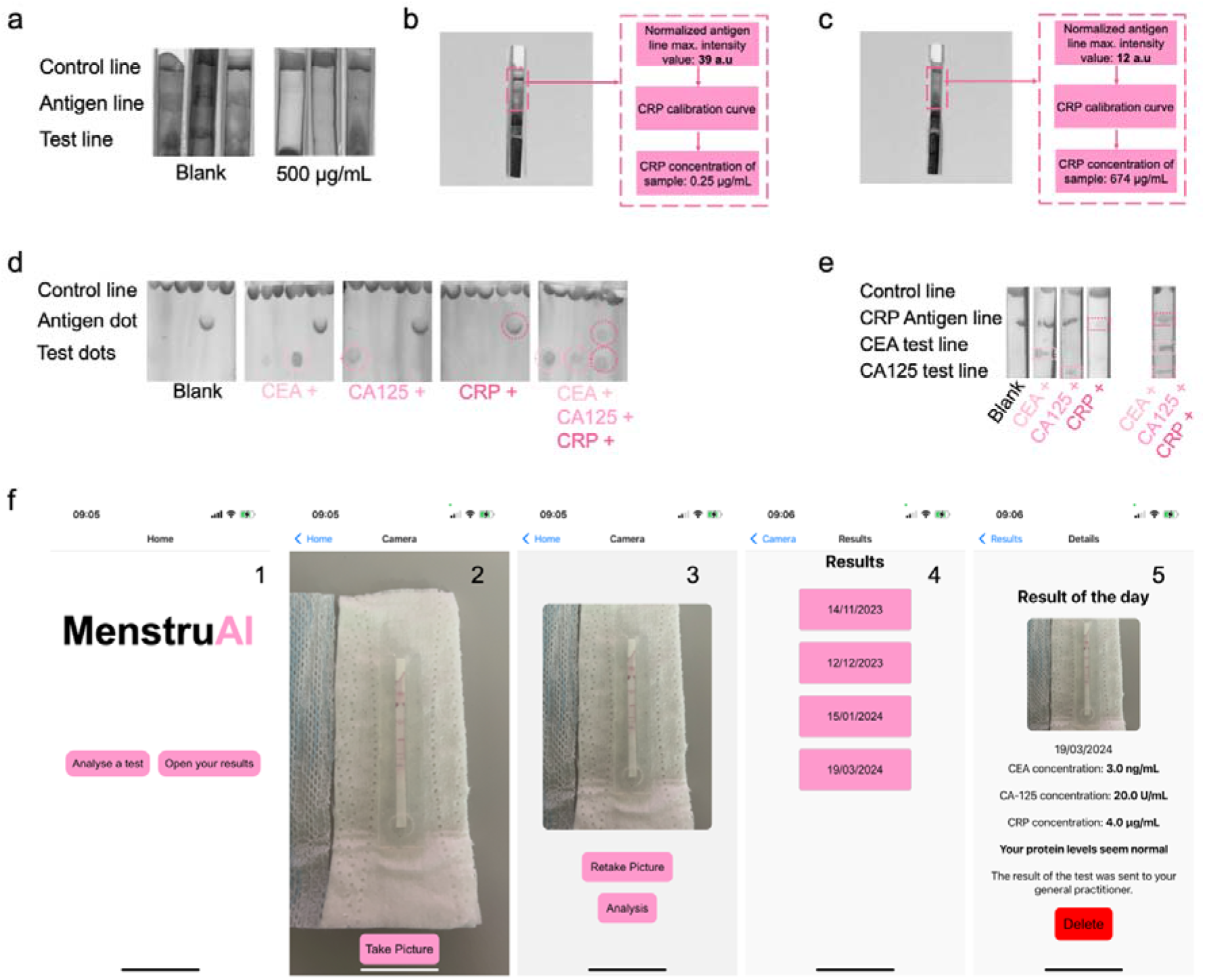
a) Example of LFA strips obtained using unprocessed whole menstruation blood for CRP concentration measurement. b) Detection of CRP in unprocessed whole menstruation blood using ML-assisted readout analysis. c) Detection of CRP in spiked unprocessed whole menstruation blood using ML-assisted readout analysis. d) Multiplexing LFA using a dot-based design. e) Multiplexing LFA using a line-based design. f) Representation of the developed smartphone application for test analysis (1: home screen of MenstruAI app. 2: camera screen to acquire an image of the test. 3: validation screen and picture analysis. 4: results screen with precedent test results folders. 5: test result folder with analysis of the image acquired and interpretation of the results). Panels show representative data from tests performed independently at least three times.

Additionally, multiplexed paper-based tests were designed to incorporate the results of the three biomarkers into a single platform. The designs used either a dot-based (Fig 6d) or line-based (Fig 6e) readout and enabled concurrent detection of at least three biomarkers (CEA, CA-125, CRP) with minimal cross-reaction. Finally, we demonstrate direct on-pad multiplex biomarker analysis using the software framework (Fig 5) integrated into a smartphone app, allowing ease of use, automatic analysis, and accessibility of the user’s data (Fig 6f). The app contains an intuitive and simple user interface, allowing the user to take a picture of the test through the app using the smartphone camera, and analysis of the test either immediately or later. The analysis report with the respective CEA, CA-125 and CRP menstruation blood levels, along with an interpretation based on the normal clinical range, can then be found alongside precedent reports in the result folder of the app (Fig 6f).

## CONCLUSIONS

In this work, we present a fully integrated on-pad detection platform for the semi-quantitative analysis of a panel of biomarkers relevant for inflammation and infection, cancer, and endometriosis, directly in menstruation blood. This work directly promotes the use of a sample often seen as waste and increases the accessibility of women to cutting-edge technologies centered on women’s health. Additionally, this first-of-its-kind point-of-care device features an innovative design that enables fluidic manipulation and multiplexed electronic-free readout, allowing for completely passive (without active human intervention) and reproducible testing in resource-constraint settings. This work presents two key innovations, namely the direct analysis of unprocessed, undiluted menstruation blood, and most importantly, fluid-control mechanisms, that enable accurate dosing of menstruation blood on the sample pad and prevent overflowing and thus invalidation of the test results. The machine-learning assisted analysis of the LFA assists the user in data interpretation and long-term tracking of biomarkers. Due to the low-barrier characteristics of this platform, we expect significant user adoption, and potentially earlier detection of healthy complications. Importantly, the platform in its current form is not intended as a diagnostic replacing clinically established tests, but rather to alert users of potential health issues and enable close-meshed, cost-effective health monitoring. In the future, we foresee that the biomarker panel could be extended depending on the user’s interest and disease prevalence, potentially also including sexually transmitted disease (STD) biomarkers, etc. The proposed menstruation blood-based multiplexed detection platform could have a transformative impact on women’s health by offering a non-invasive, cost-effective, and barrier-free approach to health monitoring in both developed and developing countries, thus contributing to healthcare democratization.

## MATERIALS AND METHODS

### Materials

Sodium citrate dihydrate HOC(COONa)(CH_2_COONa)_2_·2H_2_O, HAuCl_4_, Phosphate-buffered saline, bovine serum albumin, sucrose C_12_H_22_O_11_, tween-20, NaH_2_PO_4_·H_2_O, Na_2_HPO_4_, Na_2_B_4_O_7_·10H_2_O, H_3_BO_3_, polyvinylpyrrolidone (PVP) (C_6_H_9_NO)_n_ were purchased from Sigma-Aldrich. Anti-CRP detection and capture monoclonal antibodies were purchased from Abcam inc. (anti-CRP C2 & anti-CRP C6). Anti-CEA detection and capture monoclonal antibodies were purchased from Fitzgerald industries international (anti-CEA F & anti-CEA G). Anti-CA-125 detection and capture monoclonal antibodies (Anti-CA-125 X325 & Anti-CA-125 X306) were purchased from Lubioscience.

Goat anti-mouse IgG were purchased from Lubioscience. CRP from human ascites were purchased from Sigma-Aldrich, and CEA proteins were purchased from Fitzgerald industries international. CA-125 human protein was purchased from Sigma Aldrich. CRP free human serum was purchased from Lubioscience, and human serum was purchased from Sigma-Aldrich. Citrate-phosphate-Dextrose anticoagulant solution was purchased from Sigma-Aldrich. All buffers and reagent solutions were prepared using distilled water or MiliQ water (>18.2Ø.cm, Milipore). The sample, conjugate, and absorbent pads, as well as nitrocellulose membranes and adhesive backing cards were obtained as part of a LFA assembly kit purchased from Nanocomposix. Additionally, Fusion 5 blood separation membrane was purchase from Cityva. Ecoflex near-clear 00-31 was purchased from Smooth-On. Sanitary pads were purchased from Always.

### Human venous and menstruation blood samples

The collection of venous (BASEC No. 2016-00816) and menstruation blood (BASEC No. 2023-00562) was approved by the cantonal ethics commission of the Canton of St. Gallen, Switzerland. Human menstruation blood samples were obtained from healthy volunteers on day 1 and 2 of menses, donated to a gynecologist at the Cantonal Hospital St. Gallen and handed over to the research lab on the same day in an anonymous way (not transferring any donor information).

### Apparatus

A Hidex sense 425-301 plate reader was used for the absorbance measurement and verification of nanoparticles conjugation. A Prusa extrusion 3D printer was used for 3D printing of parts. An iPhone 13 along with ImageJ software analysis were used for quantitative evaluation of the assay’s results.

### Reagents preparation

Phosphate buffer (0.01M, pH 7.4) was made using Na_2_HPO_4_ and NaH_2_PO_4_.H_2_O in MiliQ water. Borate buffer (100mM, pH9) was made using Na_2_B_4_O_7_·10H_2_O and H_3_BO_3_ in miliQ water. The sample pad buffer consisted of a solution of PBS containing 0.5% (w/v) BSA and 1% (v/v) tween-20. Finally, the conjugate pad buffer in which nanoparticles were resuspended after conjugation consisted of a solution of PBS containing 5% (w/v) sucrose, 1% (w/v) BSA and 0.5% (v/v) tween-20.

### Gold nanoparticle synthesis

To synthesize gold nanoparticles, a 60 mL solution of 2.2 mM of sodium citrate dihydrate was prepared in 3 Neck round bottom glass flask and brought to boiling point. 400 µL of a 25 mM HAuCl_4_ was added afterwards, resulting in seed nanoparticles of approximately 6 to 10 nm diameter after 15 min. Afterwards, a growth step was realized adding 400 µL of a 60 mM sodium citrate dihydrate solution the mix, followed by 400 µL of a 25 mM HAuCl_4_ solution. After cooling down, gold nanoparticles of approximately 20 nm diameter were fully synthesized. To obtain larger diameter nanoparticles, additional growth steps were completed until the nanoparticles reached the desired diameter.

### Conjugation of antibodies to gold nanoparticles

To prepare gold nanoparticle conjugates, the desired detection antibodies were passively conjugated to the surface of the nanoparticles from non-covalent interaction. Briefly, two solutions of 40 nm AuNP at pH9 were respectively mixed with a 300 µg/mL solution of anti-CEA antibody diluted in MiliQ water and a 350 µg/mL solution of anti-CA-125 antibody diluted in MiliQ water followed by the addition of 1% BSA in MiliQ water for the obtention of anti-CEA-AuNP and anti-CA-125-AuNP conjugates. Similarly, a solution of 20 nm AuNP at pH8 was mixed with a 200 µg/mL solution of anti-CRP antibody diluted in MiliQ water followed by the addition of 1% BSA in MiliQ water. After incubation for 30 min, the 40 nm AuNP-based mixtures and 20nm AuNP-based mixtures were respectively centrifuged at 3500 and 10,000 rpm and resuspended in the conjugate pad buffer.

### Lateral flow assay preparation

The LFA sensor for individual biomarker detection consists of four components: a sample pad (blood filtration pad or not), a nitrocellulose membrane, a conjugate pad, and an absorbent pad. The sample pad was pre-treated with a sample pad buffer to assist in the fluid migration. The desired conjugated nanoparticles in a conjugate pad buffer were deposited on the conjugate pad. The absorbent pad was left untreated. For the CEA-LFA, a line of a 1 mg/mL anti-CEA capture antibody solution in phosphate buffer was deposited on the NC membrane to act as the test-line, and a 1.0 mg/mL goat anti-mouse IgG solution in phosphate buffer was deposited to act as the control line. For the CA-125-LFA, a line of a 0.75 mg/mL anti-CA-125 capture antibody solution in phosphate buffer was deposited on the NC membrane to act as the test-line, and a 1.0 mg/mL goat anti-mouse IgG solution in phosphate buffer was deposited to act as the control line. For the CRP LFA, a line of a 0.75 mg/mL anti-CRP capture antibody solution in phosphate buffer was deposited on the NC membrane to act as the test-line. An extra line of a 1.0 mg/mL anti-CRP capture antibody solution in phosphate buffer was deposited, followed by the addition of a 1.0 mg/mL human CRP solution deposited on top of the same line, to act as the antigen line. The control line was formed by a 1.0 mg/mL goat anti-mouse IgG in phosphate buffer. All components were dried for 1h in a 37°C drying oven, and later assembled on a plastic adhesive backing card. Each segment overlapped its neighboring one by 2 mm. Finally, individual LFA were cut to obtain regular paper-based sensors of 5 mm width and 6 cm length.

### Test procedure

Once LFA were prepared, solutions of the biomarker of interest were prepared in different matrices at various concentration. CEA, CA-125 and CRP solutions were prepared by spiking human serum and CRP-free human serum, as well as human whole venous blood containing anticoagulant, at desired concentrations. Additionally, CRP solutions were prepared by spiking human whole menstruation blood at desired concentrations. 60 to 100 µL of sample were deposited on the sample pad. Once the test was performed, images were obtained using an iPhone 13 and analyzed with ImageJ. The intensity profile of individual color channels of the LFA strip image was obtained, and the peak intensity of the lines was recorded. The intensity value was obtained by subtracting the peak value of the line from the background value. All measurements were triplicated.

### In-pad biosensor assembly

Fabrication of soft silicon casing started with the design of negative parts and molds in Fusion 360 CAD software, followed by 3D printing with an extrusion 3D printer. Ecoflex near-clear 00–31 part A and part B were mixed in 1:1 (v/v) ratio. The mix was later poured into the desired mold adding the negative parts to form the desired geometries after curing for 4H at room temperature. The negative parts were later removed manually, and biopsy punchers were used to create inlets and outlets in the silicon casing. The LFA sensors were later integrated into the silicon casing to obtain a fully assembled test platform. PVP membranes for volume control were synthesized by mixing PVP powder in methanol at various concentration depending on the desired retention time to achieve. Once the mix was fully dissolved, the solution was drop casted onto circular shaped Teflon mold and left to dry for 1H. Once ready, the PVP membranes were directly added to the silicon casing. A commercial sanitary pad was used as a proof of concept. The three layers composing the pad were separated, and the soft-silicon device was integrated below the first polypropylene sheet, acting as transfer layer for any fluid encountering the pad. The whole assembly was later used as such for experiments.

### Artificial intelligence assisted analysis using smartphone app

The smartphone-based analysis using AI has been built using three distinct machine learning models to i) detect the readout zone ii) classify the type of readout and iii) perform peak segmentation on the intensity profile of the LFA strip to correlate the test line intensity to the biomarker concentration. The workflow consists of automatically detecting the readout zone on the LFA. The readout zone is the zone where the output of the LFA is visible thanks to AuNP-based colorful lines. Once the zone is automatically detected, the output is analyzed and classified depending on the type of line visible. A model is used to classify the readout zone in three distinct categories (control line only; test and control lines; test, antigen, and control lines visible). The pixel intensity profile of the region is then obtained and automatically processed with a bespoke model, to extract the value of the test line intensity peak that later correlates with the biomarker concentration using pre-determined calibration curves.

First, a dataset containing 549 images of individual strips of LFA was built, incorporating LFA for the detection of CEA, CA-125 and CRP with different fluids (phosphate buffered saline, human serum, and blood). The dataset has been manually labelled to draw a bounding box containing the AuNP-based colorful lines on each individual strip. The dataset was divided into training, testing and validation sets representing 70%, 20% and 10% respectively. Transfer learning has been leveraged, using a pre-trained yoloV8 convolutional neural network^36^ model to learn the detection and segmentation of the readout zone using 50 epochs and a batch size of 10.

In a second time, a dataset for the classification of the readout has been built. 549 images of individual readout zones were cropped out from the images of single LFA strips and separated depending on the readout (control line only; test and control lines; test, antigen, and control lines visible). The dataset was divided into training, testing and validation sets representing 70%, 15% and 15% respectively. The same pre-trained yolov8 CNN model was used to learn the classification of a given LFA readout using 50 epochs and a batch size of 5.

Finally, the automatic peak segmentation was performed using a fully connected layer model. To do so, intensity profiles were manually extracted for each of the 549 individual strips. They were manually labelled, attributing a probability 1 to the pixels displaying a colorful line on the LFA and a probability 0 to the pixels consisting of the background region on the LFA. The labelled profiles were subsequently separated in two: profiles containing test line and control line-based peaks, and profiles containing test line, antigen line and control line-based peaks. For the two types, the dataset was divided into training and testing sets representing 80% and 20% respectively, and two identical FCL models were trained using the pixel position and the corresponding intensity as input for supervised learning of the probability of the pixel to fall within a peak region or a background region. The two models were built from an input layer of 512 nodes, a hidden layer of 256 nodes and an output layer of 1 node. Dropout of 0.5 was used in-between layers, as well as a ReLu activation function. The sigmoid activation function was used for the output layer. The models were trained using binary cross entropy as a loss function over 100 epochs and batch size of 10. A binary response was finally obtained from the probability output by the model using a threshold value of 0.5.

The smartphone-based application has been coded using JavaScript and React. The open-source framework React Native has been used alongside Expo Go client app to develop an application available on both iOS and Android platforms.

## Data Availability

All data produced in the present study are available upon reasonable request to the authors

## ACKNOWLEDGEMENTS

We thank the anonymous menstruation blood donors for their participation in this study, Stefan Bjelajac for his help in creating the image database, and Oscar Cipolato for proofreading and providing feedback on the manuscript. We acknowledge funding by ETH Zurich and in parts by the Swiss National Science Foundation (Eccellenza grant no. 181290, I.K.H).

