## Supplementary material for "A Wearable In-pad Diagnostic for the Detection of Disease Biomarkers in Menstruation Blood": Fig S

+41 (0)58 765 7153

**Establishment of LFA for CEA, CA-125 and CRP in serum and whole blood**

LFA sensors were designed for the detection and semi-quantification of individual biomarkers in relevant biofluids and clinically relevant detection windows. Careful optimization of the LFA sensors was performed to achieve detection and semi-quantification in i) human serum, and ii) unprocessed human whole blood.

**Direct measurement of CEA level in human serum**

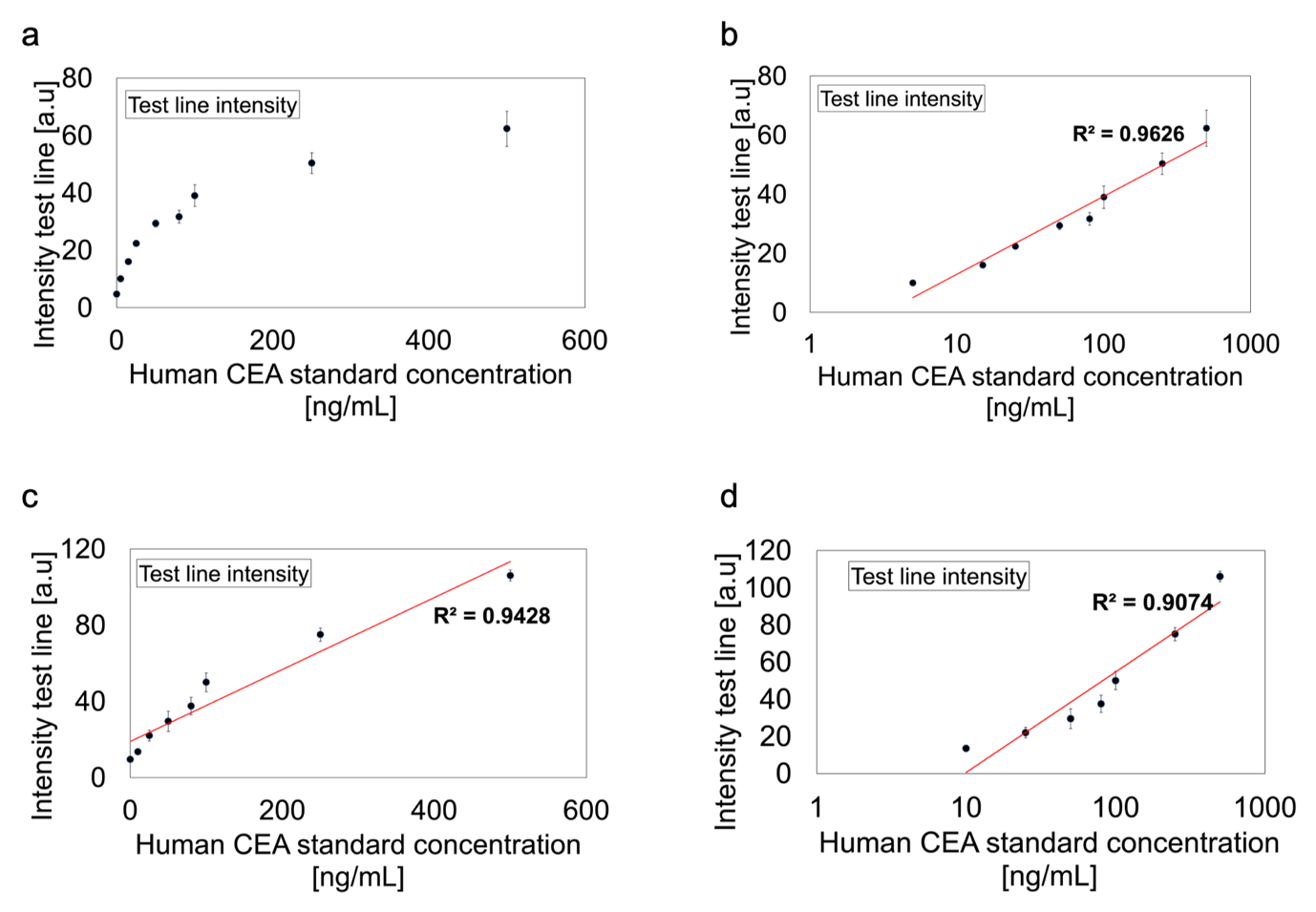

**Figure S1:** Detection and quantification of CEA using CEA LFA with human serum. a) A logarithmic response curve was observed with increasing concentration of CEA between 0 and 500 ng/mL. b) Considering a log scale, a linear response curve can be obtained on the entire range of concentration with a coefficient R^2^ of 0.96

**Direct measurement of CRP level in human serum and human whole blood**

The full CRP concentration range (0 to 500 µg/mL) is rarely measurable due to the hook effect. At concentrations above 10 µg/mL (Fig. S2 a,c), the test line intensity decreased while the CRP concentration increased, rendering interpretation of the test challenging due to false negatives. Using a sandwich assay-based line, a linear detection window was only obtained for concentrations of CRP in serum and blood between 0 and 10 µg/mL (Fig S2 a,b,c). This demonstrates the necessity to introduce a competitive assay-based line. In both human serum and human whole blood, the signal generated by this line allowed interpretation of the test results on the full CRP concentration range (0 to 500 µg/mL) without generating false negatives (Fig S2 d).

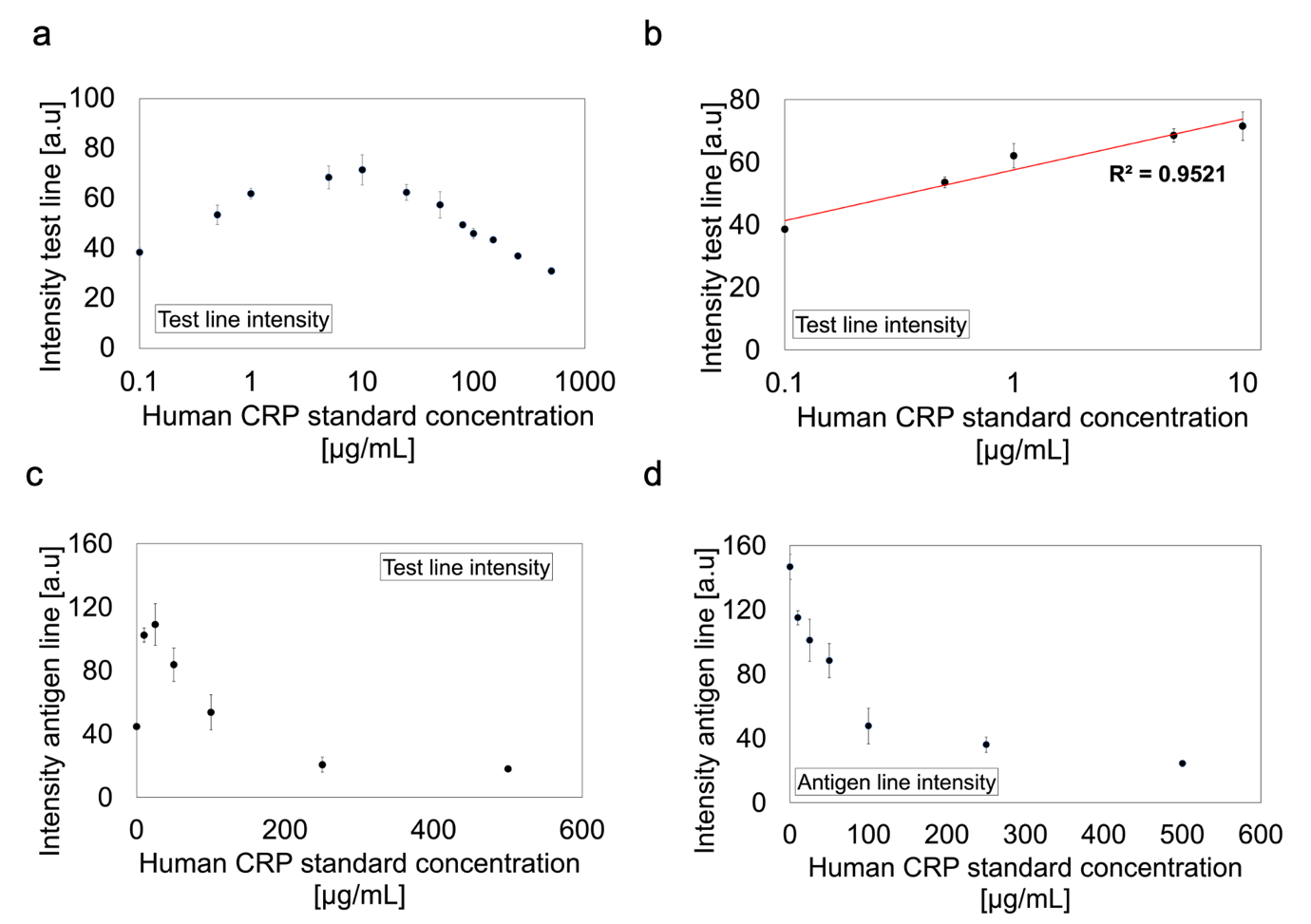

**Figure S2:** Detection and quantification of CRP using CRP LFA with a) human serum and b) human whole blood. a) The test line intensity response with increasing CRP concentration showed the occurrence of the hook effect at concentration of CRP in human serum above 10 µg/mL. b) A linear detection response was obtained between 0.1 and 10 µg/mL using the test line response with a coefficient R^2^ of 0.95. c) The test line intensity response with increasing CRP concentration showed the occurrence of the hook effect at concentration of CRP in human whole blood above 10 µg/mL. d) The competitive assay-based line intensity response showed no occurrence of the hook effect with increasing concentration of CRP on the entire concentration range (0 to 500 µg/mL).
